# Using random forests to understand unrecognized progression to late-stage CKD, a case-control study

**DOI:** 10.1101/2021.10.14.21264915

**Authors:** Christopher Hane, Stephan Dunning, Jeff McPheeters, David Mosely, Jennifer St. Clair Russell, Donna Spencer

## Abstract

**Background and objectives:** Patients with undiagnosed CKD are at increased risk of suboptimal dialysis initiation and therefore reduced access to home dialysis and transplantation as well as poor outcomes. Improved understanding of how patients remain undiagnosed is important to determine better intervention strategies.

**Design, setting, participants, and measurements:** A retrospective, matched, case-control analysis of 1,535,053 patients was performed to identify factors differentiating 4 patient types: unrecognized late-stage CKD, recognized late-stage CKD, early-stage CKD and a control group without CKD. The sample included patients with commercial insurance, Medicare Advantage, and Medicare fee-for service coverage. Patient demographics, comorbidities, health care utilization, and prescription use were analyzed using random forests to determine the most salient features discriminating the types. Models were built using all four types, as well as pairwise for each type versus the unrecognized late-stage type.

**Results:** Area under the curve for the three pairwise models (unrecognized late-stage vs recognized late-stage; unrecognized late-stage vs early-stage; unrecognized late-stage vs no CKD) were 82%, 68% and 82%.

**Conclusions:** The lower performance of the unrecognized late-stage vs early-stage model indicates a greater similarity of these two patient groups. The unrecognized late-stage CKD group is not simply avoiding or unable to get care in a manner distinguishable from the early-stage group. New outreach for CKD to undiagnosed or undetected, insured patients should look more closely at patient sets that are like diagnosed early-stage CKD patients.

## Introduction

Over 37 million U.S. adults have chronic kidney disease (CKD), and approximately 800,000 patients live with end-stage kidney disease (ESKD) (1,2). The majority of individuals with CKD are unaware of their disease, putting them at risk of receiving inappropriate care and not receiving services to delay CKD progression (2). Individuals with late referral to nephrology care may have reduced access to transplantation and home dialysis, suboptimal dialysis initiation, poor clinical outcomes, increased mortality, and higher health care costs (1). Improved identification of CKD is essential for educating patients about the impacts of kidney disease and supporting them in an effort to delay the need for kidney replacement therapies.

The primary aim of this study was to determine prognostic factors that differentiate between patients with known progression from early to late-stage CKD versus patients who have no evidence of earlier stage CKD when they first appear with late-stage CKD or ESKD. While CKD can progress very quickly in some patients, it is often a disease that worsens over many years. With the noted lack of disease awareness and under-diagnosis in earlier stages, we make the reasonable assumption that individuals with undetected (or undiagnosed) CKD prior to a late-stage diagnosis have progressed from an earlier stage, albeit “unrecognized.” Thus, we characterize this class of individuals in our study as, having *unrecognized progression to late-stage CKD or ESKD*.

This study builds on prior research in two ways. First, many studies of unrecognized kidney disease have focused on individuals who “crash into dialysis” or have a “suboptimal” dialysis start (3-9). In this study, our primary population of interest includes not just patients with ESKD without prior CKD diagnosis but also patients with late-stage CKD (G4 or 5) who lack prior recognition. This study is one of the first to use an integrated set of commercial claims, Medicare Advantage (MA) and Medicare fee-for-service (FFS) claims.

Second, the study uses a non-parametric machine learning method, random forests, to create classification models of four patient groups. Random forests are collections of randomized trees that partition the data set to minimize classification errors (10,11).

## Methods

### Data

This study used de-identified administrative claims data from the OptumLabs® Data Warehouse (OLDW), with linked socioeconomic status information (12). The database contains longitudinal health information on enrollees representing a mixture of ages, ethnicities and geographical regions across the United States. The claims data in OLDW includes medical and pharmacy claims and enrollment records for commercial, MA and Medicare FFS enrollees.

The Medicare FFS claims files held by OptumLabs, a certified CMS qualified entity, were approved for research re-use by CMS. This study is exempt from IRB review and obtained an exemption from the New England Institutional Review Board (NEIRB).

### Design

Subjects were required to have at least 12 months of continuous enrollment in any combination of plans with both medical and pharmacy coverage between 2011 and 2017 and to be at least 40 years old. For Medicare beneficiaries, Parts A, B, and D coverage were required. Individuals dually eligible for Medicare FFS and Medicaid were included because Medicare is the primary payer for their services.

Four classes of subjects were identified for the analysis: 1) Patients with late-stage CKD or ESKD without prior CKD recognition, 2) patients with late-stage CKD or ESKD with prior CKD recognition, 3) other patients with CKD, and 4) patients without CKD. Patients with CKD were identified at their most severe stage of disease using claims data and multiple diagnosis dates. Patient data was collected from claims in the one year prior to the index date.

Lab claims were not used in the identification rules. Diagnoses in the claims were confirmed using ICD-9 and -10 codes on at least one inpatient claim in the primary position or two claims in any position on separate dates at least 30 days apart within a 1-year period. All code sets are found in the supplement. We refer to this logic of inpatient and multiple outpatient visits as confirmation of the diagnoses; this is not a confirmation by a lab test.

There were two inclusion criteria for unrecognized progression to late-stage CKD and ESKD (Class 1). One was patients had late-stage CKD (defined as CKD stages G4 and G5) or ESKD. The patient index date was the earliest late-stage CKD or ESKD diagnosis date. The second criterion was the absence of evidence for earlier-stage/unspecified CKD more than 30 days and up to 12 months before the index date. One percent of patients were in class 1.

Class 2 patients had the same criteria for confirmed diagnosis as Class 1 but also had to have a diagnosis for CKD (earlier stage or unspecified) in the 12 months prior to index date, not including the 30 days prior to index date. One percent of patients were in class 2.

Class 3 patients had a diagnosis for earlier-stage CKD (CKD G1-G3, or unspecified) without evidence of a late-stage CKD/ESKD diagnosis. The patient’s index date was randomly set to the service date of a CKD claim on or after their confirmed CKD diagnosis. The study intentionally used a random CKD encounter date as the index date because using the last early-stage date showed a strong bias to patients near end of life. 13% of patients were in class 3.

Class 4 were the non-CKD patients who do not meet the requirements of classes 1-3. Their index date was set to a random encounter date. Note, it was possible that some patients in this class had limited evidence of CKD that did not meet our claims-based definition for a CKD diagnosis. 85% of patients were in class 4.

In all four classes, all subjects were required to have 12 months of continuous enrollment before the index date and to not have evidence of pre-index date kidney transplant (see supplement for codes). Individuals who had any evidence of hospice care or who had long-term care stays amounting to over 90 days during the pre-index period were excluded.

#### Matching

Because risk of CKD varies with age, we matched subjects across the four classes in the training data using 5-year age bands to avoid spuriously detecting an age-related correlation between the outcome classes. The study also matched on Census region, Medicare and Medicaid dual-eligibility status, gender, and index year.

We determined the number of unique matching strata as the product of the number of unique values of each of the matching variables. Each subject in the training set was then assigned to a stratum from 1 to 1,679 (the total number of strata in the data set) based on their value of each of the matching variables. Within each stratum, the maximum number of possible Class 1 patients was determined that met the matching ratios. The number of Class 1 patients then determined the number of matched patients in the other classes. The specific patients meeting the matched count were randomly selected.

Table 1 summarizes the final matching counts. The match rate refers to the fraction of the original training data that was matched within a class, whereas the cohort fraction refers to the proportion of the final training data set represented by a class. The final matched training data set included 1,535,053 subjects. Sensitivity analysis showed no meaningful deviation in match rates by matching variables (data not shown).

**Table 1.** Matching Results by ML Class.

| Class | Matched Subjects | Cohort Fraction | Match Rate (%) |
| --- | --- | --- | --- |
| 1 | 150,553 | 9.8 | 91.5 |
| 2 | 180,100 | 11.7 | 92.2 |
| 3 | 451,635 | 29.4 | 23.4 |
| 4 | 752,765 | 49.0 | 5.9 |
| Total | 1,535,053 | 100.0 | 10.3 |

#### Analytic Methods

We used random forests to create models that separate the subjects into the four classes. Four separate models were evaluated: Class 1 vs. 2, Class 1 vs. 3, Class 1 vs. 4, and a multinomial model with all four class labels. This set of models was chosen to evaluate the importance of model features in differentiating each class from Class 1 as well as their global importance.

Random forests compute a local frequency of class outcomes using recursive partitioning of the data. As the name suggests, the forest is a collection of decision trees where each tree starts with an independent random sample with replacement of the training data. This randomization of the data for each tree improves model generalization. Each tree recursively selects one feature that improves the classification the most then splits the data set into two new data sets as children of that node. The features available to the algorithm at each node are also randomly selected. This helps uncover correlated features and reduces run time.

The analysis included a comprehensive set of features measured during the 12 months prior to a subject’s index date:

- **Demographics:** These features included age, gender, race/ethnicity, index year, region, insurance type, Medicaid/Medicare dual eligibility status, and whether a subject had a change in insurance type during the pre-index year.
- **Comorbidities:** A subject’s medical diagnoses were summarized by Quan Charlson Comorbidity Index score (13). Additionally, binary flags for specific disease conditions as categorized by the Agency for Healthcare Research and Quality (AHRQ) Clinical Classifications Software (CCS) were included (14). CCS codes were customized to remove the diagnosis codes used in class definitions (i.e., CKD codes to remove perfect predictors), and select CCS categories were split into two or more groups to isolate kidney-related conditions and other conditions of interest, resulting in a total of 308 classifications.
- **Health care utilization:** The analysis included counts of primary care visits, ambulatory (outpatient and office) visits, ER visits, inpatient stays, long term care visits, nephrologist and other specialist visits, kidney and A1c tests, medical nutritional therapy visits, and total health care costs. The subject’s Charlson score was used to standardize the subject’s total medical costs into a percentile value per Charlson group (0, 1, 2-3, 4+), resulting in a cost burden variable that varied from one to 100.
- **Pharmacotherapy utilization:** Total days’ supply was calculated for each of over 300 medication classes defined the American Hospital Formulary Service (AHFS)(15). Two variables capturing pharmacy benefits (type of pharmacy plan and deductible for medications) were included.

All continuous features were winsorized at their 98^th^ percentile, then divided by that percentile to have a range of zero to one. This uniformity in range assists in interpreting the variable importances from the multi-variate analysis (16). The models had 675 to 692 features with the difference due to rare features missing in some models.

The models were fit using a random forest as implemented in the R package ranger, version 0.12.1 run within R version 4.0.2 (10, 17). All forests were computed with 1,000 trees with a max depth of 8 and at least 500 subjects per node. The project took advantage of ranger’s impurity corrected variable importance that adjusts the importance for bias in the variable selection common in other implementations.

The statistics used for the evaluation were the classification error and area under the receiver operating curve (AUC) as computed on the out-of-bag (OOB) data as is standard for random forests. Out-of-bag refers to the subjects not chosen in the sampled data of one tree

## Results

Table 2 shows the post-matching descriptive profile of each class. The values for features used in matching agree across the classes. Race varies across the classes from 17% black in Class 2 to 10% black in Class 4; and 71% white in Class 2 to 77% white in class 4. Race was not used as a matching variable to understand its role as a predictor. Note that the data is matched on Medicaid dual eligibility status which may act as a surrogate for poverty.

**Table 2.** Cohort Profile.

| Characteristic | Statistic | Class 1<br>N=150,553 | Class 2<br>N=180,100 | Class 3<br>N=451,635 | Class 4<br>N=752,765 |
| --- | --- | --- | --- | --- | --- |
| Gender |  |  |  |  |  |
| Female | N | 82,529 | 98,745 | 247,578 | 412,645 |
|  | % | 54.82 | 54.83 | 54.82 | 54.82 |
| Male | N | 68,024 | 81,355 | 204,057 | 340,120 |
|  | % | 45.18 | 45.17 | 45.18 | 45.18 |
| Age | N | 150,553 | 180,100 | 451,635 | 752,765 |
|  | MEAN | 74.47 | 74.50 | 74.46 | 74.34 |
|  | MEDIAN | 77 | 77 | 77 | 77 |
|  | STD | 9.21 | 9.15 | 9.20 | 9.22 |
| Age category |  |  |  |  |  |
| 40-49 | N | 3,056 | 3,533 | 9,162 | 15,280 |
|  | % | 2.03 | 1.96 | 2.03 | 2.03 |
| 50-64 | N | 17,058 | 20,298 | 51,162 | 85,290 |
|  | % | 11.33 | 11.27 | 11.33 | 11.33 |
| 65+ | N | 130,439 | 156,269 | 391,311 | 652,195 |
|  | % | 86.64 | 86.77 | 86.64 | 86.64 |
| Race |  |  |  |  |  |
| White | N | 106,310 | 125,640 | 335,709 | 582,860 |
|  | % | 70.61 | 69.76 | 74.33 | 77.43 |
| Black | N | 23,951 | 30,823 | 62,046 | 72,704 |
|  | % | 15.91 | 17.11 | 13.74 | 9.66 |
| Hispanic | N | 12,860 | 14,904 | 32,876 | 59,000 |
|  | % | 8.54 | 8.28 | 7.28 | 7.84 |
| Asian | N | 5,367 | 6,196 | 15,516 | 27,758 |
|  | % | 3.56 | 3.44 | 3.44 | 3.69 |
| Unknown | N | 2,065 | 2,537 | 5,488 | 10,443 |
|  | % | 1.37 | 1.41 | 1.22 | 1.39 |
| Region |  |  |  |  |  |
| Northeast | N | 25,789 | 30,813 | 77,367 | 128,945 |
|  | % | 17.13 | 17.11 | 17.13 | 17.13 |
| South | N | 63,425 | 75,992 | 190,275 | 317,125 |
|  | % | 42.13 | 42.19 | 42.13 | 42.13 |
| Midwest | N | 37,425 | 44,784 | 112,275 | 187,125 |
|  | % | 24.86 | 24.87 | 24.86 | 24.86 |
| West | N | 23,727 | 28,326 | 71,181 | 118,635 |
|  | % | 15.76 | 15.73 | 15.76 | 15.76 |
| Unknown | N | 187 | 185 | 537 | 935 |
|  | % | 0.12 | 0.10 | 0.12 | 0.12 |
| Insurance type |  |  |  |  |  |
| Commercial | N | 2,546 | 2,573 | 8,301 | 18,958 |
|  | % | 1.69 | 1.43 | 1.84 | 2.52 |
| Medicare Advantage | N | 15,784 | 19,143 | 47,124 | 73,314 |
|  | % | 10.48 | 10.63 | 10.43 | 9.74 |
| FFS/FFS+commercial | N | 132,223 | 158,384 | 396,210 | 660,493 |
|  | % | 87.83 | 87.94 | 87.73 | 87.74 |
| Baseline coverage change | N | 1,620 | 1,856 | 5,319 | 8,654 |
|  | % | 1.08 | 1.03 | 1.18 | 1.15 |
| Index dual eligibility (among FFS) | N | 55,601 | 66,577 | 166,261 | 274,425 |
|  | % | 42.05 | 42.04 | 41.96 | 41.55 |
| RX deductible |  |  |  |  |  |
| Non-zero RX deductible | N | 74,808 | 87,536 | 222,470 | 385,976 |
|  | % | 49.69 | 48.60 | 49.26 | 51.27 |
| Unavailable | N | 3,356 | 4,302 | 11,806 | 26,492 |
|  | % | 2.23 | 2.39 | 2.61 | 3.52 |
|  | % | 12.14 | 12.12 | 12.15 | 12.14 |

For each model, the study recorded the variable importance of all variables, the OOB prediction error, and OOB area under the curve (AUC).

Table 3 presents the statistics for each model. Note that the Class 1 prevalence reflects the matching ratio for each class described earlier (e.g., for Class 2, prevalence = 1/(1+ 1.2) or 0.455). All pairwise model AUC values were over 0.8 except for the Class 1 vs. 3 model. That model’s AUC below 0.7 indicates that the model was not able to discriminate as well as the other models. The ‘all’ model generates 4 class probabilities for each subject. This model has no global AUC value because the AUC measure is only computable for a binary outcome. In its place, the table reports the AUC scores for each class measured to an outcome of that class label vs any other label. For example, the Class 1 AUC in the ‘all’ model is computed using only the Class 1 probabilities with an outcome of Class 1 or not.

**Table 3.** Model Results for Each Class Outcome and Overall.

| Model (Outcome) | Prevalence of Class 1 | OOB Prediction Error | Training Error | AUC |
| --- | --- | --- | --- | --- |
| Class 1 vs. 2 | 0.455 | 0.195 | 0.263 | 0.817 |
| Class 1 vs. 3 | 0.250 | 0.179 | 0.242 | 0.677 |
| Class 1 vs. 4 | 0.167 | 0.116 | 0.136 | 0.818 |
| All | 0.098 | 0.372 | 0.098 | 0.694 |
|  |  |  | 0.106 | 0.875 |
|  |  |  | 0.291 | 0.701 |
|  |  |  | 0.240 | 0.846 |

Random forests compute a value for the variable importance that measures the improvement in solution quality as the variable (i.e., model feature) is selected to partition the data set. This study divided the raw importance by the total importance for all variables in each model. This yielded a percentage of total importance for each variable, a more easily comparable value across the models.

Table 4 shows the 41 variables that explained at least 1% of the total importance in at least one of the four models. The rows are ordered by the sum of the scaled importances. Only seven variables met the 1% threshold in all three of the class-specific models: nephrology visits, nephrology unique providers, Charlson score, age, blood panel testing, urinary creatinine testing, and loop diuretics. This group of variables separate Class 1 from the other three classes.

**Table 4.** Variables Explaining at Least 1% of Total Importance by ML Model.

| Variable | Fraction of total importance explained in each model |  |  |  |
| --- | --- | --- | --- | --- |
|  | Class 1<br>vs. 2 | Class 1<br>vs. 3 | Class 1<br>vs. 4 | All |
| Nephrology, unique providers | 13.04 | 4.63 | 7.05 | 8.46 |
|  | Class 1<br>vs. 2 | Class 1<br>vs. 3 | Class 1<br>vs. 4 | All |
| Nephrology visits | 14.32 | 5.82 | 6.64 | 8.83 |
| Charlson score | 7.51 | 11.48 | 4.34 | 10.69 |
| Blood panel | 8.25 | 4.05 | 1.94 | 5.84 |
| Primary care, unique providers | 1.87 | 4.39 | 0.15 | 0.82 |
| Primary care visits | 2.81 | 5.40 | 0.20 | 1.28 |
| Other diseases of kidney and ureters | 2.61 | 0.37 | 8.21 | 5.36 |
| Acute and unspecified renal failure | 1.50 | 0.73 | 9.08 | 5.12 |
| Loop diuretics | 1.10 | 3.19 | 6.70 | 3.03 |
| Age | 1.45 | 5.21 | 3.44 | 1.53 |
| Urinary creatinine lab test | 4.65 | 2.29 | 1.01 | 3.06 |
| Diabetes with kidney-related complications | 3.29 | 0.59 | 3.55 | 3.11 |
| Insulins | 0.36 | 1.56 | 3.89 | 1.66 |
| Congestive heart failure, nonhypertensive | 0.50 | 0.56 | 3.33 | 1.82 |
| Direct vasodilators | 0.96 | 0.99 | 2.47 | 1.19 |
| Outpatient hospital visits | 2.64 | 2.35 | 0.04 | 0.53 |
| Diabetes with other complications | 0.40 | 0.41 | 2.71 | 1.67 |
| Office visits | 2.25 | 2.17 | 0.08 | 0.51 |
| Other non-specialist visits | 1.68 | 2.12 | 0.19 | 0.94 |
| Deficiency and other anemia | 1.37 | 0.12 | 1.52 | 1.76 |
| Beta-adrenergic blocking agents | 0.70 | 0.27 | 2.02 | 1.66 |
| Other encounter days | 1.44 | 1.05 | 0.71 | 1.33 |
| Hypertension | 0.94 | 0.84 | 0.92 | 1.84 |
| Antigout agents | 0.73 | 0.15 | 2.29 | 1.21 |
| Fluid and electrolyte disorders | 1.19 | 0.21 | 1.25 | 1.69 |
| Dihydropyridines | 0.61 | 0.72 | 1.76 | 1.07 |
| Devices | 0.25 | 0.48 | 2.22 | 1.05 |
| Urinary albumin lab test | 0.90 | 0.68 | 0.70 | 1.48 |
| Hyperlipidemia | 0.70 | 2.28 | 0.03 | 0.48 |
| HgbA1c lab test | 0.80 | 0.30 | 1.00 | 1.31 |
| Inpatient stays | 0.82 | 1.42 | 0.36 | 0.75 |
| Diabetes with no complications | 0.36 | 0.10 | 1.46 | 1.42 |
| Genitourinary symptoms and ill-defined conditions – kidney complications | 1.86 | 0.12 | 0.47 | 0.89 |
| Nephritis, nephrosis, renal sclerosis | 0.79 | 0.03 | 1.48 | 0.84 |
| Other specialist visits | 0.79 | 1.95 | 0.05 | 0.11 |
| Vitamin D | 1.64 | 0.03 | 0.47 | 0.71 |
| Unknown | 0.46 | 1.36 | 0.07 | 0.20 |
| Sulfonylureas | 0.03 | 0.16 | 1.00 | 0.51 |
|  | Class 1<br>vs. 2 | Class 1<br>vs. 3 | Class 1<br>vs. 4 | All |
| Immunizations and screening for infectious disease | 0.22 | 1.36 | 0.01 | 0.01 |
| Other screening for suspected conditions (not mental disorders or infectious disease) | 0.06 | 1.22 | 0.06 | 0.00 |

The variable importances show that the Class 1 vs. 3 model is different as its variable importance scores are often outliers in each row of table 4. This model had the lowest AUC score which indicates that it lacked the ability to differentiate the subjects in this pair of classes. The Class 1 vs. 3 model had six unique variables meeting the 1% explained threshold: screenings, immunizations, unknown provider visits, other specialist visits, IP stays, and hyperlipidemia. The appearance of screenings and immunizations and other non-specific features as the most important in this model indicate that there was not a clear set of clinical markers to separate Class 1 from Class 3.

While Class 4 is a population without evidence of CKD in claims, it would be a mistake to label the class as “healthy”. Table 4 shows that Class 1 is different from Class 4 primarily by acute kidney disease and other kidney disease (9% and 8% of total importance) followed by nephrology utilization as measured by unique nephrology providers and visits (5% and 6%). Primary care visits did not play an important role in differentiating Class 1 vs Class 4, nor did other utilization measures (all < 0.2%).

Figures 1 and 2 display the means of comorbidity and utilization feature distributions which show that class 1 and 3 are often ranked 2^nd^ and 3^rd^ respectively. Class 2 has highest prevalence and utilizations and Class 4 the lowest.

**Figure 1.**
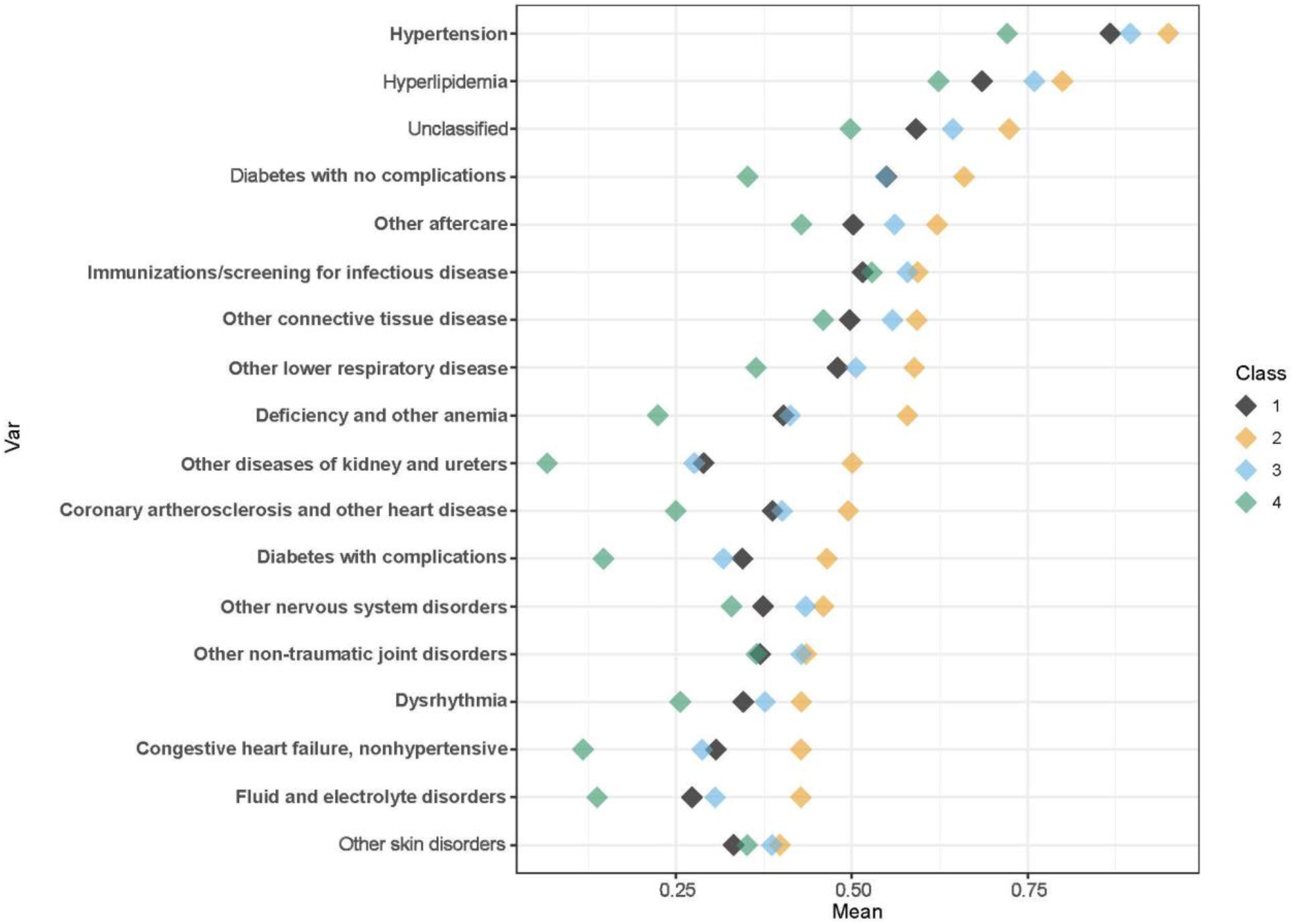
Prevalence of Comorbidity by Class

**Figure 2.**
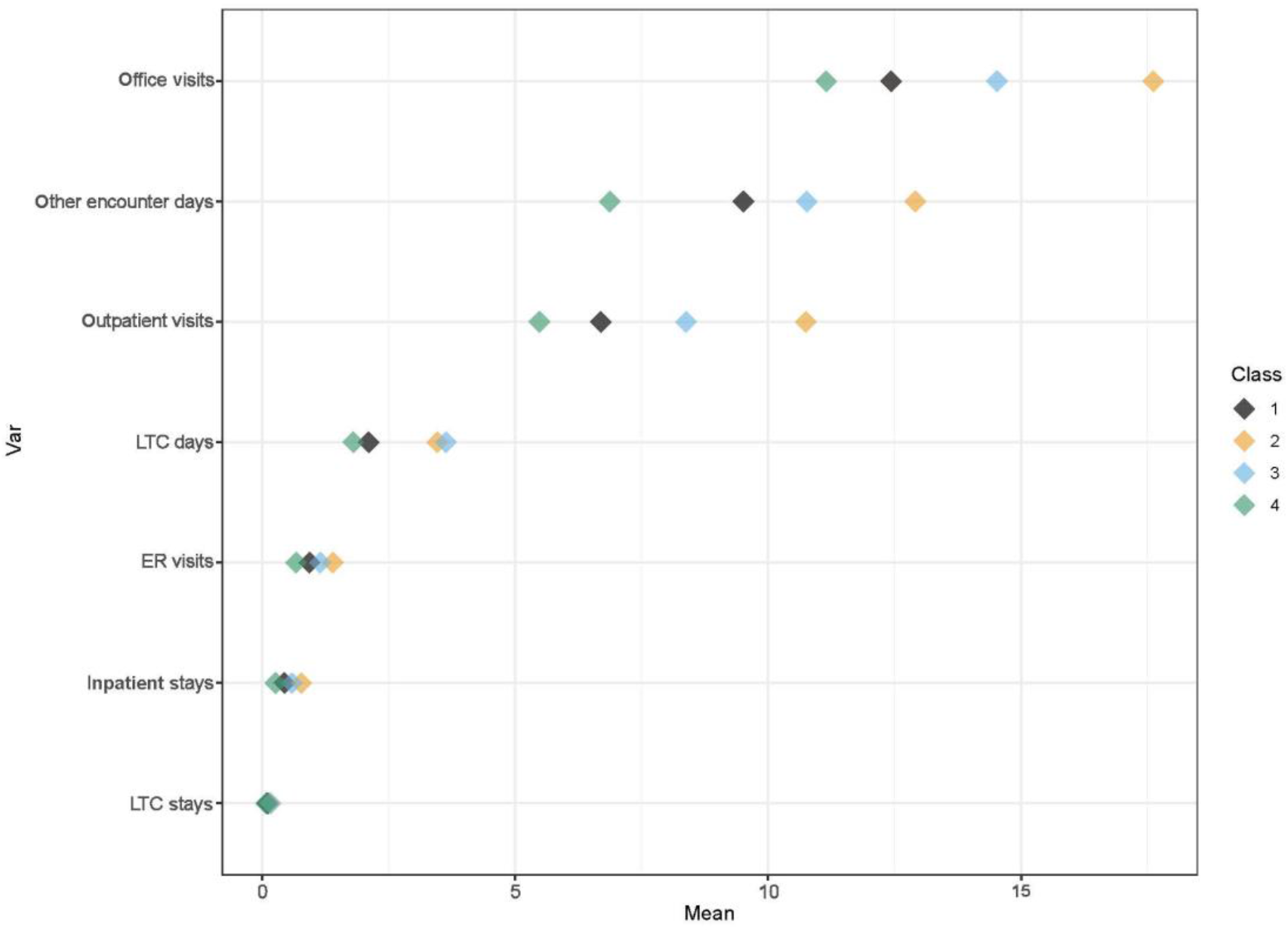
Mean Utilization by Class

## Discussion

To close the substantial gap in under-diagnosis of CKD, we need to better understand the differences between patients who are diagnosed earlier in the course of the disease versus not. That was the ultimate goal of this study. Past research has shown that older age, male gender, increased comorbidity burden, diabetes, cardiovascular disease (e.g., congestive heart failure), and hypertension are associated with late referral to nephrologist care and suboptimal dialysis initiation (18-21). The lack of recent nephrologist care also has been found to be a significant predictor of suboptimal dialysis starts (21).

This study broadened the definition of the group of people in the previous literature from initiating dialysis to unrecognized late-stage CKD or ESKD to understand if similar factors are also important as we evaluate patients upstream from ESKD. This study also subset the patient classes to determine variation in degree of association between the medical factor and patient class.

We confirm that nephrology visits, comorbidity burden, diabetes and hypertension play a role in discriminating the patient class (Table 4). The degree of importance of these factors varies across the pairs of patient classes – nephrology being the most important and hypertension the least.

Class 1 vs Class 4 variable importance was highest for acute kidney failure, other diseases of the kidney, nephrology visits and loop diuretics. These factors indicate that while Class 1 is unrecognized CKD, they do have a history of kidney problems. Clinicians managing patients with diabetes and other risk factors for CKD should be monitoring for kidney problems such as nephrotic syndrome, which has been shown to be associated with higher risks of ESKD, venous thromboembolism (VTE), atherosclerotic cardiovascular events, and heart failure hospitalizations (22).

The importance of loop diuretics across all models may indicate that patients are being treated for edema prior to diagnosis of CKD, or that the treatment is occurring without a full investigation of kidney function. Further research on this question may be helpful.

An initial hypothesis was that people with unrecognized progression to late-stage CKD/ESKD (Class 1) may be ‘care avoiding’ or have significant barriers to accessing care, and for these reasons, they are unrecognized. Figure 1 shows the utilization rates by class do not support this, nor does Figure 2 showing the prevalence of the CCS comorbidities. While there may be a subset of class 1 that fits that hypothesis, the figures and the low AUC for Class 1 vs Class 3 support that Class 1 looks most like Class 3.

The lower AUC performance (67.7%) in the case of unrecognized late-stage vs early stage CKD indicates a greater similarity of these two patient groups. The unrecognized late-stage CKD group does not have low healthcare utilization in a manner distinguishable from the early-stage group. New outreach for CKD to undiagnosed or undetected, insured patients should look more closely at patient sets that are like early-stage CKD patients.

Our analysis shows there are both distinct differences and similarities between people with unrecognized progression to late-stage CKD/ESKD and the other groups of patients that could be used to help improve risk models for more effectively using limited resources to monitor populations for enabling earlier detection and intervention of chronic kidney disease.

## Data Availability

While de-identified data was used, the data use agreements require that researchers apply directly to CMS and Optum Life Sciences for access to the data.

## Acknowledgments

The authors wish to thank Joseph Vassalotti, MD and Ashby Andrews (National Kidney Foundation); Kevin Plosser, MBA and Rahul Dhawan, MD (Optum Kidney Services); Jon Friedman, MD, FAST (Optum Medical Benefit Management); Kevin Larsen, MD (OptumLabs); Anil Chandraker, MD (Brigham and Women’s Hospital); Josef Coresh, MD, PhD and Raquel Greer, MD, MHS (Johns Hopkins University); John Cuddeback, MD, PhD (American Medical Group Association); Alex Liang, MD (Dallas Nephrology Associates); Connie Rhee, MD (University of California, Irvine, School of Medicine); and Wendy St. Peter (University of Minnesota) for their input and feedback on this research as members of the Steering Committee or Technical Expert Panel for this project. The authors also acknowledge Lillian Hang, MBA, MPH and Pamela Morin, MBA, both of OptumLabs, for their programming support and Molly Diethelm, PMP of OptumLabs for her project management support in this work. Select results from this study were presented at the 2021 National Kidney Foundation Virtual Spring Clinical Meetings, April 6-10, 2021.

## References

1. Chronic Kidney Disease in the United States, 2021 [Internet]. 2021 Available from: https://www.cdc.gov/kidneydisease/publications-resources/ckd-national-facts.html Accessed May 4, 2021.

2. United States Renal Data System. US renal data system 2019 annual data report: Epidemiology of kidney disease in the United States. [Internet] 2019. Available from: https://www.usrds.org/media/2371/2019-executive-summary.pdf Assessed May 4, 2021.

3. Chiu K, Alam A, Iqbal S: Predictors of suboptimal and crash initiation of dialysis at two tertiary care centers: Crash and suboptimal dialysis initiation. Hemodial Int 16: S39–S46, 2012

4. Heaf J, Heiro M, Petersons A, Vernere B, Povlsen JV, Sørensen AB, Clyne N, Bumblyte I, Zilinskiene A, Randers E, Løkkegaard N, Ots-Rosenberg M, Kjellevold S, Kampmann JD, Rogland B, Lagreid I, Heimburger O, Lindholm B: Suboptimal dialysis initiation is associated with comorbidities and uraemia progression rate but not with estimated glomerular filtration rate. Clinical Kidney Journal 14: 933–942, 2021

5. Molnar AO, Hiremath S, Brown PA, Akbari A: Risk factors for unplanned and crash dialysis starts: a protocol for a systematic review and meta-analysis. Syst Rev 5: 117, 2016

6. Middleton RJ, Foley RN, Hegarty J, Cheung CM, McElduff P, Gibson JM, Kalra PA, O’Donoghue DJ, New JP: The unrecognized prevalence of chronic kidney disease in diabetes. Nephrology Dialysis Transplantation 21: 88–92, 2006

7. Go AS, Yang J, Tan TC, Cabrera CS, Stefansson BV, Greasley PJ, Ordonez JD: Contemporary rates and predictors of fast progression of chronic kidney disease in adults with and without diabetes mellitus. BMC Nephrol 19: 146, 2018

8. Onuigbo MAC: Syndrome of rapid-onset end-stage renal disease: a new unrecognized pattern of CKD progression to ESRD. Renal Failure 32: 954–958, 2010

9. Krolewski AS, Skupien J, Rossing P, Warram JH: Fast renal decline to end-stage renal disease: an unrecognized feature of nephropathy in diabetes. Kidney International 91: 1300–1311, 2017

10. Wright MN, Ziegler A: ranger : A Fast Implementation of Random Forests for High Dimensional Data in C++ and R. J Stat Soft [Internet] 77: 2017 Available from: http://www.jstatsoft.org/v77/i01/ [cited 2020 Dec 3]

11. Biau G, Scornet E: A random forest guided tour. TEST 25: 197–227, 2016

12. Wallace PJ, Shah ND, Dennen T, Bleicher PA, Crown WH: Optum Labs: Building A Novel Node In The Learning Health Care System. Health Affairs 33: 1187–1194, 2014

13. Quan H, Li B, Couris CM, Fushimi K, Graham P, Hider P, Januel J-M, Sundararajan V: Updating and Validating the Charlson Comorbidity Index and Score for Risk Adjustment in Hospital Discharge Abstracts Using Data From 6 Countries. American Journal of Epidemiology 173: 676–682, 2011

14. Clinical Classification Software (CCS) for ICD-9-CM. Agency for Healthcare Research and Quality, [Internet]. Available from: http://www.hcup-us.ahrq.gov/toolssoftware/ccs/ccs.jsp [cited 2020 Dec 12]

15. AHFS Therapeutic Classification [Internet]. Available from: https://www.ashp.org:443/Products-and-Services/Database-Licensing-and-Integration/AHFS-Therapeutic-Classification [cited 2020 Dec 12]

16. Hastie T, Tibshirani R, Friedman JH: The elements of statistical learning: data mining, inference, and prediction. 2nd ed. New York, NY, Springer.

17. R: The R Project for Statistical Computing [Internet]. Available from: https://www.r-project.org/ [cited 2020 Dec 3]

18. Singhal R, Hux JE, Alibhai SMH, Oliver MJ: Inadequate predialysis care and mortality after initiation of renal replacement therapy. Kidney International 86: 399–406, 2014

19. Arulkumaran N, Navaratnarajah A, Pillay C, Brown W, Duncan N, McLean A, Taube D, Brown EA: Causes and risk factors for acute dialysis initiation among patients with end-stage kidney disease—a large retrospective observational cohort study. Clinical kidney journal. 12: 550–558, 2019

20. Navaneethan, SD., Aloudat, S, Singh, S: A systematic review of patient and health system characteristics associated with late referral in chronic kidney disease. BMC Nephrology 9: 1–8, 2008

21. Hassan R, Akbari A, Brown PA, Hiremath S, Brimble KS, Molnar AO: Risk Factors for Unplanned Dialysis Initiation: A Systematic Review of the Literature. Can J Kidney Health Dis 6: 2054358119831684, 2019

22. Go AS, Tan TC, Chertow GM, Ordonez JD, Fan D, Law D, Yankulin L, Wojcicki JM, Zheng S, Chen KK, Khoshniat-Rad F, Yang J, Parikh RV: Primary Nephrotic Syndrome and Risks of ESKD, Cardiovascular Events, and Death: The Kaiser Permanente Nephrotic Syndrome Study. JASN ASN.2020111583, 2021

