## Supplement for "Using random forests to understand unrecognized progression to late-stage CKD, a case-control study"

### Appendix

#### Table 1. Class Definitions for ML Analysis

| Class | | Core Inclusion/Exclusion Criteria |
|---|---|---|
| 1 | Patients with late-stage CKD (defined as CKD stages G4 and G5) or ESKD ***without*** prior CKD recognition within the claims data | • Evidence of late-stage CKD or ESKD between January 1, 2012 and December 31, 2017 (identification period): Late-stage CKD and ESKD were identified based on diagnosis codes (see Appendix). Patients were required to have at least one non-lab inpatient claim with a late-stage CKD or ESKD diagnosis (primary position) or two non-lab claims with a diagnosis (any position) on separate dates at least 30 days apart within a 1-year period. The ***index date*** was set as the earliest evidence of late-stage CKD or ESKD (i.e., the earliest diagnosis code) during the identification period.<br><br>• The absence of one non-lab inpatient claim (primary position) or two non-lab claims (any position) at least 30 days apart with a diagnosis code for early-stage or unspecified CKD (i.e., unrecognized) in the 12 months prior to the index date. If an enrollee had 1+ non-lab claim for early-stage or unspecified CKD in the 30-day period before the index date but no non-lab claim(s) earlier within the 12-month period prior to the index date, the enrollee was considered "unrecognized." See Appendix for codes used in defining early-stage and unspecified CKD. |
| 2 | Patients with late-stage CKD or ESKD ***with*** prior recognition of CKD within the claims data | • Evidence of late-stage CKD or ESKD (same as above) between January 1, 2012 and December 31, 2017. As with Class 1, the ***index date*** was set at the earliest evidence of late-stage CKD or ESKD during the identification period.<br><br>• At least one non-lab inpatient claim (primary position) or two non-lab claims (any position) at least 30 days apart with a diagnosis code for early-stage or unspecified CKD (i.e., recognized) in the 12 months prior to the index date. If an enrollee had 1+ non-lab claim for early-stage or unspecified CKD in the 30-day period before the index date but no non-lab claim(s) earlier within the 12-month period prior to the index date, the enrollee was not considered "recognized." |
| 3 | Other patients with CKD (i.e., patients not in Classes 1 or 2 above; includes patients with recognized CKD stages G1-3 or stage not specified) | • Failure to meet the above criteria for late-stage CKD/ESKD.<br><br>• Evidence of early-stage or unspecified CKD between January 1, 2012 to December 31, 2017. Enrollees were required to have at least one non-lab inpatient claim with an early-stage or unspecified CKD diagnosis (primary position) or two non-lab claims with a diagnosis (any position) at least 30 days apart within a 1-year period. The ***index date*** was set as a random service date for CKD during the identification period that came on or after the enrollee meeting the definition of early-stage or unspecified CKD and occurred in an enrollment window during which the early-stage/unspecified CKD definition criteria were met. |
| 4 | Patients without any history of CKD[1] | • Failure to meet the above criteria for late-stage CKD/ESKD/other CKD.<br><br>• The index date was set as a random service date during the identification period. |

[1] Note: It was possible for some patients to fall in different cohorts at different points in time. For example, it was possible that patients in Cohort 4 had evidence of CKD during a time that we did not observe in the data. Further, within the study data and timeframe, we could have observed a patient in Cohort 3 at one point in time and then Cohort 2 at another. For the purpose of this study, patients were assigned to the highest cohort (e.g., 2 instead of 3) in such cases.
